# An online, group Acceptance and Commitment Therapy is acceptable to stroke survivors: A qualitative interview study

**DOI:** 10.1101/2024.09.05.24313129

**Authors:** Hannah Foote, Audrey Bowen, Sarah Cotterill, Emma Patchwood

**Affiliations:** Manchester Centre for Health Psychology, School of Health Sciences, Geoffrey Jefferson Brain Research Centre, MAHSC, University of Manchester, UK; Centre for Biostatistics, School of Health Sciences, University of Manchester, UK

**Author notes:** **corresponding author** Dr Hannah Foote.

## Abstract

Mental health difficulties are common post-stroke and developing support for psychological adjustment is a research priority. Wellbeing After Stroke (WAterS) is a nine-week, online, group-based Acceptance and Commitment Therapy (ACT)-informed intervention, delivered by trained third-sector practitioners, supervised by a clinical neuropsychologist. This study aimed to explore the acceptability of WAterS from the stroke survivors ‘ perspective.

Semi-structured interviews were conducted with twelve stroke survivors who received WAterS. The interview schedule was informed by theorised components of acceptability, including understanding, burden and perception of effectiveness. The data were analysed inductively and deductively using Template Analysis.

Six qualitative themes were generated. Results indicate the intervention was mostly understandable and participants were able to engage with ACT and apply it to life. Online delivery reduced burden in accessing the intervention, and was acceptable when supported by live facilitation and a physical handbook. Group cohesion and understanding was facilitated by practitioners. The social aspect of the group was beneficial. Attending WAterS supported some participants to seek further support; others were left feeling unsupported when the intervention ended.

Stroke survivors valued attending an online, group ACT-informed intervention, delivered by practitioners. This is a promising avenue in increasing the reach of interventions to support wellbeing.

## Introduction

Mental health issues occur frequently post-stroke ^1–3^ and support for psychological difficulties is the number one research priority for life after stroke ^4^. Acceptance and Commitment Therapy (ACT) ^5^ is a trans-diagnostic, third-wave, cognitive behavioural therapy with growing evidence for its use post-stroke ^6–10^.

The Wellbeing After Stroke (WAterS) feasibility study ^11^ co-developed and demonstrated the feasibility of a protocolised, nine-week, online ACT-informed intervention delivered to small groups of stroke survivors experiencing self-reported distress and difficulties adjusting.

Intervention groups were led by trained practitioners employed by Stroke Association (a national voluntary organisation), who received weekly supervision from a clinical neuropsychologist. A detailed intervention and training description is available^11^.

The Theoretical Framework of Acceptability (TFA)^12,13^ defines acceptability as “the extent to which people delivering or receiving a healthcare intervention consider it to be appropriate, based on anticipated or experienced cognitive and emotional responses to the intervention”^13^ and posits seven components of acceptability. These include intervention coherence (the extent to which the participants understand the intervention), burden and perceived effectiveness (participants ’ perceptions as to whether the intervention will achieve its purpose). All seven TFA components and their definitions are included in supplemental material S3. Examining acceptability can inform intervention modifications to improve the design of future research and implementation ^14,15^ and can be usefully explored via qualitative means ^15,16^.

In the extant literature on ACT for stroke survivors there has been scant investigation of acceptability. One study of ACT delivered to groups of people with acquired brain injury found high acceptability as measured by a satisfaction rating scale ^8^. One qualitative study has explored stroke survivors ’ experiences of receiving an ACT-informed intervention ^17^, but this was not explicitly focused on acceptability.

This study is the first to explore the acceptability of an ACT-informed intervention from the perspective of the stroke survivors who have received it, using a Theoretical Framework of Acceptability lens.

## Materials and Methods

Research ethics approval was secured from the University of Manchester (ref 2021-11134-18220). This study is reported using the COnsolidated criteria for REporting Qualitative research (COREQ) checklist ^18^ (see Supplemental material S1).

This qualitative study used semi-structured, one-to-one interviews and took a ‘limited realist ’ position ^19^, to recognise the subjectivity of the participants and researchers, while drawing on theory and assuming that findings have the potential for wider relevance.

Participants self-referred to the WAterS study, following advertising via Stroke Association mailing lists and social media. The eligibility criteria were:

1. Adults (at least 18 years old)
2. At least 4 months post-stroke (no upper limit)
3. Self-report as having unmet needs in terms of psychological adjustment to stroke and psychological distress
4. Sufficient English language to engage in groups and complete measures
5. Ability to engage in remote group interventions.

Taking part in this interview study was a voluntary addition to the host WAterS feasibility study^11^. Recruitment was contemporaneous with the host study and online via Zoom.Participants could opt to have an informal carer present during the interview to support with technology access and understanding. Following recruitment, demographic information and assessment data were gathered – these are summarised here and reported in full elsewhere^11^.

### Study materials

The interview schedule was developed by the authors in collaboration with a Patient, Carer and Public Involvement (PCPI) advisory group. The schedule was informed by the Theoretical Framework of Acceptability ^13^, and addressed each of the seven theorised components of acceptability (see Supplemental material S2 for full interview schedule).

### Data collection and processing

The interviews were conducted by female authors HF (PhD student) and EP (Postdoctoral Researcher), both with previous experience of interviewing for qualitative research. The researchers had no prior relationship with the participants, but had contact with them as part of the host WAterS feasibility study^11^. They took place as soon as possible following the end of the WAterS intervention sessions to support the participants in recalling their experience. Interviews occurred via Zoom, in a private location. If an informal carer was present they were asked not to contribute their views. Interviews were recorded (audio and video) and securely stored. Field notes were taken as necessary. The recordings were transcribed verbatim and checked for accuracy.

### Data analysis

Data were managed using NVivo software. Findings were thematically analysed using Template Analysis ^20^, which allows both inductive and deductive analysis and use of *a priori* themes. The analytic process included eight procedural steps ^20,21^:

1. Identified *a priori* themes: the seven TFA components
2. Read through all transcripts to familiarise with the data
3. Coded the data by *a priori* and themes inductively generated by researchers
4. Produced initial template of themes, grouped as either top-level or sub-themes
5. Applied initial template to full data set and re-defined, modified and collapsed top-level and sub-themes
6. Quality checks were carried out, and the template further revised:
  a. Two authors separately coded one transcript according to the template, then compared and discussed codes to facilitate reflections on the analysis
  b. Feedback was given on initial data analysis by the WAterS PCPI group
7. Template of themes finalised
8. Final template applied to full data set.

## Results

All 12 participants who took part in the WAterS intervention groups accepted the invitation to be interviewed for this study, in October—December 2021, within nine days of completing the intervention. The mean length of interview was 60 minutes (39–90 minutes). One participant had an informal carer (their spouse) present during the interview.

A summary of participant information relevant to this paper is provided here, with full baseline data reported in the WAterS feasibility study paper^11^. Seven males and five females were recruited. 10 participants were white, one black and one Asian. The mean age of participants was 53.7, ranging from 34 to 76. Five participants had received the UK statutory level of education, three had A Levels and four had University degrees. The mean number of months post-stroke was 25 (range 5–90). Participants had mild-to-moderate cognitive and communication difficulties and self-reported as having unmet needs in terms of psychological adjustment to stroke and psychological distress.

## Findings

The final template used for analysis comprised of six study specific themes, developed by the researchers to best represent the data to answer the research objective. Table 1 shows the six themes and how they map onto relevant *a priori* TFA themes (also see Supplemental material S3 for the iterative development of templates used during analysis).

**Table 1:** The six study specific themes and examples of how these relate to the TFA components.

| Theme | Relevant TFA components |
| --- | --- |
| Engaging with ACT and applying it to life | *Intervention coherence* – understanding increased ability to apply strategies to life<br>*Perceived effectiveness* – participants provided examples of successfully applying strategies to their lives |
| Seeking community and support | *Affective attitude* – participants had both positive and negative feelings about the groups they were in<br>*Ethicality* – for some, contributions from other group members did not fit with their individual values<br>*Perceived effectiveness* – learning from others in the group increased perceptions of effectiveness<br>*Self-efficacy* – participants’ confidence in taking part was impacted on by how comfortable they felt in the group |
| Adaptations are important to support accessibility | *Burden* – some found aspects of the intervention effortful to access, despite adaptations. Reminders reduced effort in remembering to attend. Online intervention reduced effort in accessing sessions<br>*Intervention coherence* – understanding was supported by provision of adapted materials<br>*Perceived effectiveness* – difficulties in accessing content reduced perceptions of effectiveness |
| Intervention structure needs to be clear and account for burden | *Intervention coherence* – a reduced pace may have increased understanding for some<br>*Opportunity costs* – participants gave up time when sessions over-ran, and the time needed to complete home practice was difficult for some<br>*Perceived effectiveness* – some felt that restructuring of the sessions would increase effectiveness |
| Facilitation supported learning and group cohesion | *Affective attitude* – participants generally felt positive about the practitioners<br>*Intervention coherence* – facilitation supported understanding of the content<br>*Perceived effectiveness* – facilitation supported participant's to contribute and to apply the material to their lives |
| Moving on: what next after WAtErS? | *Affective attitude* – some participants felt disappointed by the intervention coming to an end<br>*Perceived effectiveness* – for some, the group ended prior to them being ready, potentially reducing its effectiveness. For others, WAtErS successfully supported them to seek out appropriate follow up support. |

The six study specific themes generated were: ‘Engaging with ACT and applying it to life ’, ‘Seeking community and support ’, ‘Adaptations are important to support accessibility ’, ‘Intervention structure needs to be clear and account for burden ’, ‘Facilitation supported learning and group cohesion ’, and ‘Moving on: what next after WAterS? ’.

### Engaging with ACT and applying it to life

While there were some challenges, most participants were able to engage with the WAterS group sessions and apply the ACT-informed skills they learnt to their lives. A number of participants discussed a change in identity post-stroke, with varying levels of acceptance. The intervention addresses these changes directly and for some this was emotionally challenging, particularly those earlier post-stroke. One participant left some sessions early when they felt challenged (but returned in subsequent sessions) and another reported finding it uncomfortable to witness others ‘ strong emotions. Other participants were at a place in their individual stroke journey where they were more able to reflect and apply the ACT strategies: “I ‘m not the person I was before. As to whether I ’ll ever be the person I was before, who knows? I could be a new and improved one […] It’s helped me stop and […] be more present, because I know there might be some horrible moments, but this moment will have something in it that’s good” [ID022].

The stroke survivors understood that the WAterS intervention included many different strategies and activities, and that they could choose to use the ones which worked for them: “There were about four or five different strategies given to us […] some hit, some miss, but that’s the nature, when you try things” [ID002].

Levels of understanding impacted on participants ‘ ability to apply the strategies. For one participant, this was particularly the case with home practice tasks, which were difficult to understand outside of the sessions: “I suffer a little bit with thoughts because everything’s still up in the air […] it’s just constantly circling round […]. It was hard to remember or understand the book [client handbook], what I was actually supposed to be doing” [ID012].

However, many stroke survivors were able to give examples of how they had taken the strategies learnt on the course and effectively applied them to their own lives: “I ’m sitting there thinking all kinds of things and getting a bit anxious, and I thought […] I ’ll use the stop. That moment of stopping, breathing” [ID006]. One participant had been avoiding an activity which was linked to where they had been when their stroke occurred. Over the course of the WAterS intervention, they were able to overcome this and re-engage in the activity.

Some participants misunderstood the purpose of session activities and/or commented on wanting further explanation and discussion as to the purpose of certain activities: “I think you need to spend more time on going over what the function of it is or why do it […] and how it might be useful” [ID006].

### Seeking community and support

Many participants volunteered to join WAterS due to a lack of support available to them, saying “I ’ll do anything to try and improve the way I feel about myself” [ID009]. Some participants had a stroke during the COVID-19 pandemic, and this was the first time they had connected with other stroke survivors. The shared experience of stroke was important to many participants: “Knowing that everybody who had been on the course had experienced stroke […] I think that facilitated a sense of unity” [ID002].

Most participants found forming connections online successful, they had become accustomed to this due to the pandemic and found they were “still able to bond with people” [ID022]. One participant found being online actively positive as they found it easier to open up via this format.

For many participants, the group provided the opportunity for the normalisation of post-stroke experiences and the opportunity to learn from each other’s experiences: “We were learning coping strategies from the other survivors and other ways of looking at things […] but also we were offering something to someone else” [ID018].

For others, differences in circumstances (such as working status and time post-stroke) and personal ethics made it difficult to relate to the other members of their group. For example, one participant stated you “have to look at those positives” [ID009], which contrasted with another participant who wanted to share experiences of hardship. For a few participants these differences exacerbated feelings of isolation “because I was just very aware that there was a massive void of where we ’re all at.” [ID009]. Conversely, many participants felt confident to take part in their group as “we got to know each other that well” [ID004].

Participants ’ differing communication styles impacted on group dynamics. Many group members were able to adapt their communication to suit the group, for example, they gave a “five-minute offering […] and then […] tried to hang back” [ID011]. However, there were a few instances where participants felt that someone else “took over the show” [ID009].

Many participants felt that having groups of four stroke survivors worked well and “it didn ’t feel people were talking over each other, it felt like there was enough space for people to talk” [ID012]. Some felt that six people per groups would be beneficial as it “might have still held people together but created more variety” [ID011].

One group had a participant’s partner present, and this participant reported that the WAterS intervention had had a positive impact on their relationship. A participant from a different group also referred to the importance of including family members: “One thing […] that I think is really important is not just to have people on these courses that have had a stroke […] but their families and loved ones because they don ’t know how to manage it” [ID009].

### Adaptations for stroke are important to support accessibility

Participants felt that having a paper client handbook was beneficial, particularly for an online intervention. They received the handbook in advance of the sessions which supported them in feeling prepared, and during the intervention they used it to follow along with the facilitation, as an aide memoire, and to record notes: “Having it physically with you, having a handbook that you can hold and look at and write in […] is actually really important now. It’s far, far better […] than having something totally online” [ID006].

One participant with aphasia stated that to make the client handbook fully accessible, it would need to be adapted in collaboration with people with aphasia. However, most participants found the handbook to be understandable with the support of the practitioners: “It’s broken down and the text is big enough. It’s quite wordy but […] the ladies that ran it […] they read things out to us, so we could follow it” [ID019].

The participants were varied in how easy they found it to understand the WAterS intervention content, in particular relating to more abstract concepts and some terminology caused confusion: “It was mentioned so many times, daily noticing, mindful, mindful noticing and stuff, and […] it just got a little bit tangled up” [ID012].

Post-stroke impairments (such as pain, tinnitus and vision problems) impacted on participation in certain activities. One participant commented that the possibility of such impairments impacting on activities should be explicitly addressed during the WAterS intervention.

The remote nature of the WAterS intervention reduced burden in accessing the course, and while some participants would have preferred to meet face-to-face, the benefits of remote access were acknowledged: “If you asked me to choose, I would say definitely face to face but […] because we ‘ve got strokes and we ’ve got some people that are quite disabled, the practicalities of getting people together is probably not ideal” [ID009].

For one participant, being part of an online group reduced access to the content as it was more difficult to speak privately to a practitioner and clarify difficulties with understanding, stating “on Zoom, you don ‘t always like to ask questions that you think other people have fully understood” [ID012].

Reminder texts and emails were sent prior to each session, which reduced burden and supported some participants to remember to attend. Zoom software was reported to be easy to use: “All I had to do was to click on them [Zoom link] for the next meeting. It suited me because […] I ’m not tech savvy […] it made it very easy for me” [ID011].

Participants were asked about when post-stroke they felt this intervention would be most appropriate. There were a range of views on this, from as soon as possible to one year post-stroke. However, participants acknowledged that straight after a stroke may be too soon: “Around the six-month mark because […] the first two to three months, from my perspective, my senses were on overload” [ID022].

### Intervention structure needs to be clear and take account of burden

This theme refers to the structural aspects of the WAterS intervention, such as length of sessions and the order of activities, rather than the content of the intervention.

The participants were broadly happy with the structure of the WAterS intervention. Sessions included a 20-minute break which was necessary for some participants, but too long for others. Sessions were planned to be two hours in duration, but often over-ran by 5–30 minutes. This was acceptable for the majority of participants: “They over-ran and I think it was valid […] I ’d always have that time set aside so it didn ’t matter that it overran because it’s not as though I had anything else” [ID004].

However, giving up this additional time was difficult for others, particularly participants who were working. Similarly, the weekly home practice tasks were difficult to fit in for participants with other commitments, and/or post-stroke fatigue: “I do suffer a lot from post-stroke fatigue. So when I ’m not working, I ’m usually flat out” [ID009].

The WAterS intervention structure felt coherent to some participants: “There was the holistic side to it that worked really well. So I could see the journey. By the end of it I knew that we ’d taken a journey” [ID018]. Whereas to others the activities included in each session felt disjointed: “Going from meditation into thinking about negative emotions, it was a sudden shift of gear” [ID011].

Views on the pace of the WAterS intervention were varied. For some it felt appropriate: “It was a nice steady pace, nothing was rushed, everybody had a chance to speak” [ID012]. Others felt that there was too much content, and one participant stated that there wasn ’t always enough time to fully understand each activity and reducing the content would allow more time for discussion and for practitioner’s to further support everyone’s understanding: “I felt at times that there was too much being fed into the syllabus and it may well have benefited from being pared down slightly […] we perhaps could have spent more time addressing those issues that were raised in that particular session” [ID011]. Each session started with a recap of the previous week and one participant felt this could be reduced to allow more time at the end to review progress: “There was some interesting discussions taking place at the end of the sessions […] and […] had that […] been cut at the beginning, it would have allowed for a smoother, rather than a chopped ending.” [ID002].

### Facilitation influences learning and group cohesion

Participants broadly had a very positive attitude towards the practitioners: “They were really good, they were really considerate, really caring, really supportive and they made you feel really comfortable” [ID009].

Participants felt that the live facilitation was necessary and that learning from the handbook alone would not be sufficient. The practitioners gave context to the concepts in the handbook, supporting intervention coherence and learning: “I think using it [the handbook] only works really with the course leaders […] because reading it through beforehand […] is […] not the same as when someone’s going through it with you and then adding things that you never thought of” [ID006].

The participants appreciated it when the practitioners were familiar with the content, stating “I loved […] that they paraphrased […], they weren ’t just reading it verbatim.” [ID018]. Participants liked the way the practitioners were able to guide discussion and respond to the individual contributions of the participants: “I was very impressed with how they were able to pick up on what the participants were saying […], maybe taking it on a bit further, and then asking another question.” [ID006]

The participants generally felt comfortable with the practitioners asking questions, however, one participant had difficulty in understanding some home practice tasks and was uncomfortable when asked for feedback on this: “They ’ll [the practitioners] say […] can you tell us about […] how you ’ve gone on with the homework, and sometimes […] I wasn ’t particularly comfortable with being put on the spot.” [ID012]

There was mixed feedback about the practitioners ’ ability to keep the session to time and ensure that all stroke survivors had the opportunity to input into discussions. Some felt this was done well: “Sometimes there might be somebody who’s going on a bit too much and I thought they handled that quite well […]. Also, bringing out from you something if you weren ’t really saying very much” [ID006]. Whilst others felt this could be improved to ensure everyone’s voice was heard: “I might have handled it differently because […] there were people who were far less assertive, who were just getting a little bit pushed out” [ID011].

For one participant the amount of support offered by the practitioners, and by the WAterS intervention as a whole, was not sufficient for their needs: “I did talk once to one of the ladies […] they weren ’t unresponsive or unhelpful, it was quite vague, quite shallow.” [ID019].

There were three sessions where practitioners had planned absences and a different practitioner led the session. The participants valued being informed of this change in advance and felt that the stand-in practitioners did well and that this was preferable to missing a session: “It made a difference ’cause personalities make a difference. But it was seamless, I think” [ID006].

### Moving on: what next after WAterS?

All participants completed the full course of WAterS group sessions. Many participants had difficult feelings in relation to the ending of the intervention, despite the final few weeks of the clinical protocol including some time for reflection on this ending. Some wished the sessions were on-going: “Why is therapy […] a short few weeks […]? When sometimes, the service user […] could do with some more sessions before it ends, why is there a time limit?” [ID008]. Others found the WAterS intervention long enough, but were still left with a sense of “huh what happens now?” [ID011]. Participants commented on missing the relationships they had formed, and the structure the WAterS intervention provided to their week: “It’s a bit disappointing actually because it was nice to have that Wednesday devoted to doing that. […] I do feel there is a bit of a loss there” [ID018].

Some of the participants exchanged contact details. Others who had not done this, stated that they would have liked WAterS to suggest this to them, or to facilitate on-going informal meetings for them to join: “One thing that I ’m hoping we can do is to get a Zoom meeting of the participants on an informal basis […] that could be one of the ways that this could be moved forward” [ID011].

For some participants the WAterS intervention was effective in supporting them to seek other types of support, which were appropriate to their needs. For example, some participants had identified options for social support available to them locally: “I Zoom with the Stroke Association […] we might talk about culture stuff or things that are going on in the world that you think, actually that’s what I ’m missing […] that conversation with people, face to face” [ID004]. For one participant, WAterS had effectively supported them to return to strategies that they had previously found helpful for their wellbeing, and they were seeking avenues to continue to access these strategies: “I ’m planning to begin to get back into meditation classes. I ’ve been doing meditation online” [ID006].

Others had explored avenues for further wellbeing support but found there to be no provision available in their area. For one such participant, the WAterS intervention had raised her awareness of challenging emotions and had increased her need for support, which was not available: “So there’s a possibility for something to be created and be magnified and then that person then, me in my home, have to live with that” [ID019].

## Discussion

The ACT-informed WAterS intervention was largely acceptable and valued by the stroke survivors who opted to receive it. Most participants found the community and support they were looking for. The intervention structure was suitable for most, and accessibility was increased by the course being online, using familiar technology, the provision of a client handbook and live facilitation. Participants gave examples of integrating the intervention strategies into daily life. Many participants found the end of the course difficult. Some had sought alternative mental health or social support, however, this support was not always available, highlighting the existing gap in psychological services for stroke survivors ^22^.

There has been very little previous research into the acceptability of ACT interventions for stroke survivors. Our findings are consistent with a single-case evaluation of an ACT-informed group intervention for people with acquired brain injury, which found high acceptability based on the group’s participants (n=8) completing satisfaction questionnaires^8^. The present study adds new contributions to this evidence, as it investigates an ACT-informed intervention delivered remotely by practitioners, and it explores acceptability in an in-depth manner, guided by the Theoretical Framework of Acceptability (TFA) ^13^. In addition, ways to further develop the intervention content and increase acceptability in future applications have been identified.

To our knowledge, there has been one previous qualitative study exploring stroke survivors ’ views of an ACT intervention ^17^, which interviewed participants (n=13) following attendance at a brief, ACT-informed intervention, didactically delivered by two practitioners, one being a clinical psychologist. This study and ours share many confirmatory findings, despite our study investigating experiences of a longer intervention, not directly facilitated by a clinical psychologist and including non-didactic activity. Both studies found that participants valued being with other stroke survivors and felt that paper resources (co-produced with stroke survivors) supported learning and recall. In both studies, some participants reported difficulties in understanding and applying the concepts outside of the groups, highlighting the need for the practical application of intervention strategies to be emphasised. A recent systematic review ^23^ investigating the acceptability of remotely-delivered mental health support for stroke survivors found, in line with the present study, that satisfaction was increased by easy-to-use technology, particularly technology already owned by participants.

### Strengths and limitations

There are strengths and limitations in the use of the Theoretical Framework of Acceptability (TFA) ^13^ in the present study. The use of the TFA during data collection supported a comprehensive investigation of acceptability. However, during analysis, certain TFA components were found to be difficult to interpret, for example, the ‘ethicality ’ component is related to whether an intervention is a ‘good fit with value system ’ and this concept was challenging to define and translate into an accessible interview question. During analysis, overlaps between the TFA components became apparent. For example, ‘Affective Attitude ’ is defined as ‘how a person feels about the intervention ’, but participants ’ feelings about the intervention were inextricably linked to other components of the framework, such as whether they perceived it to be effective. Validation of the TFA is ongoing and we are yet to learn its full value in exploring acceptability. Therefore, Template Analysis was used to allow for inductive exploration of our data, alongside deductive analysis using the TFA.

No sampling was required as all stroke survivors who received the WAterS intervention consented to be interviewed in this additional study. However, we did not interview the five people who initially consented to the host WAterS feasibility study^11^ but declined the invitation to attend the intervention groups.

The profile of the participants in this study is likely to have been impacted by the recruitment strategy (as self-referral via online advertising requires motivation and skill) and the remote delivery which required internet access and a device that supported Zoom. The mean age of the participants was approximately 54, which is significantly lower than the Stroke National Audit Programme’s (SSNAP) finding of 76 ^24^. Eighty-three percent of the participants were white, which is close to the 85% reported in the national stroke audit ^24^. However, due to the small number of participants the use of a percentage must be treated with caution given the need to reduce health inequalities ^25,26^.

The interview schedule was piloted with members of the WAterS PCPI group, who were also consulted regarding preliminary results and analysis. However, the interviewers were members of the WAterS research team and this may have impacted on the participants ’ responses. Quality checks were carried out during data analysis, including independent coding to prompt discussion and reflection on analysis. Interviews were carried out within nine days of completing the intervention to support recall, so data on the longer-term acceptability of this intervention was not gathered. However, follow up interviews (four-to-six months post-intervention) have now been carried out with eight of the participants and findings appear consistent with the present study ^27^. This research indicates that the Theoretical Framework of Acceptability (TFA) may be a useful tool for investigating acceptability when evaluating interventions. Researchers should carefully consider the definitions of the framework’s components in relation to their intervention of interest.

Further research is needed to investigate who the WAterS intervention is most appropriate for, to further inform inclusion/exclusion criteria, and to reduce health inequities ^25,26^ through co-development with under-served populations, including minoritised ethnic groups and stroke survivors with more severe cognitive and language difficulties ^28^. The Wellbeing After Stroke-2 study began in October 2023 to begin to address these issues ^29^.

In conclusion, this study provides preliminary evidence that an online, group, ACT-informed intervention, delivered by trained practitioners is acceptable to the stroke survivors who opt to receive it. This study provides useful insights for future work on the development and delivery of group-based and/or remotely-delivered ACT interventions for this population, from the stroke survivors ’ perspective, including the importance of easy-to-use technology, live facilitation and the provision of physical resources, co-produced with stroke survivors.

## Supporting information

Supplemental 1 - COREQ checklist

Supplemental 2 - Interview schedule

Supplemental 3 - Iterative development of qualitative themes

## Data Availability

All data referred to in this manuscript can be requested via the corresponding author. All requests will be dealt with on a case-by-case basis.

