## Supplemental 2 - Interview schedule for "An online, group Acceptance and Commitment Therapy is acceptable to stroke survivors: A qualitative interview study"

### S2: Interview schedule

- **Introduce** myself: occupation, previous education/employment.
- The interview will be **recorded** (video & audio). Re-cap key information from PIS. Stored securely –**GDPR/data protection**. Everything you say is **confidential**, unless someone is at risk of harm/malpractice/illegal activity. Your responses will be **anonymised** as soon as possible. Just the audio part will be sent to a UoM approved company for transcription. I might make the odd note, but the recording will mean I don't need to take detailed notes.
- Explain **purpose** of the interview – looking for their feedback on how they have found the groups. Explain reason for wanting to know this.
- Time scale – beginning at developing this course. Very **early stages. Really keen to know how we could improve the course for the future**
- No right or wrong answers. This is not about your knowledge, interested in your opinions/feelings.
- Acknowledge that I am from the research team – encourage honesty and welcome all positive/negative feedback.
- The interview will be **informal**. Hopefully it will feel like a conversation.
- Acknowledge that I am from the research team – encourage **honesty** and welcome all **positive/negative** feedback.
- Make clear that we need to focus on questions during the recording, but can come back to other stuff at the end **(ground rules)**
- We can do this over more than one session if you like.
- Check **permission** to go ahead with interview. Check permission to **record** the interview.

#### Questions:

| |
|---|
| What attracted you to joining the study/sessions? Can you tell me a bit about why you wanted to join the study/sessions? *[maybe ask at the end]* |
| Before you started, how did you feel about the idea of the therapy [use participants word]?<br>Were there any reservations/concerns about taking part? How do you feel about X now? |
| How do feel about the therapy groups now?<br><br>Were there particular parts that you really liked/disliked? *[consider key components here]* |
| Was there anything you've had to give anything up to participate in the groups? |
| How easy/difficult was it to take part?<br><br>What contributed to making it easy/difficult? |
| How easy/difficult was the course to understand? (pace of info)<br><br>Any key things that helped/hindered your understanding? |

| |
|---|
| Any particularly difficult parts? |
| Can you tell me a bit about your understanding of the purpose/aims of the therapy? |
| Do you feel the course has been of benefit to you?<br><br>Any aspects that have been most/least beneficial/helpful? |
| How confident did you feel to take part/contribute?<br><br>Has this changed across the course of the groups?<br>Any particular things that impacted on your confidence? (could be stroke related or not)<br>*[could ask about confidence in completing homework here or later on with other homework questions]* |
| How do you feel about the handbook? How did you use it?<br><br>Was the handbook something you looked at before starting the groups? What impact (if any) did this have?<br>Any particularly good/bad bits? Any improvements? |
| This course was developed specifically for people who have had a stroke. How do you feel about that? How did that impact on your experience of the group? |
| How did you find the homework?*[could ask for specific info on how much they did using handbook records?]*<br><br>Too much/too little?<br>How easy/difficult was to understand?<br>Confidence in completing homework?<br>Did you do it alone/with support from anyone? |
| How did you find attending a course online? (have you previously attended online groups?)<br><br>What were the benefits of attending online?<br>What were the challenges to attending online? |
| How did you find doing this within a group?<br><br>What were the benefits?<br>What were the challenges? |
| How did you feel about the staff running your group?<br><br>How did this impact on your experience of the course?<br>When you had a back-up member of staff for a session – how was this? Impact? |
| Are there any aspects of the therapy/strategies that you will carry on with?<br><br>What will you do? How do you see X helping you?<br>How confident do you feel that you will continue with X?<br><br>Do you plan to access any further support/activities for your wellbeing? |
| How did you feel about the length of time for the sessions?<br><br>How were the breaks? Enough breaks? Long enough? |

| |
|---|
| Was the fact that the sessions were longer than planned a benefit or a cost?<br>Was the time of day appropriate? |
| How did you feel about the reminder texts/emails? |
| If anyone misses a session, could ask about experience of catching up |
| Was now a good point in your stroke journey to take part in the group? When might have been better?<br>*Can you tell me a bit about why you joined when you did?* |
| Anything you would like to tell me that I haven't asked you about? |
| Any recommendations for changes/improvements to make for future groups? |
| Is there anything else you would like to comment on? |

- Thank you for your time
- Any other questions?
- Feel free to email me if you think of anything else
