## Supplemental 3 - Iterative development of qualitative themes for "An online, group Acceptance and Commitment Therapy is acceptable to stroke survivors: A qualitative interview study"

### S3: Iterative development of the templates used for analysis

#### Initial template one (a priori themes): The Theoretical Framework of Acceptability (TFA) components[1,2]

| **Main themes (TFA components)** | ***TFA definitions*** |
|---|---|
| Affective Attitude | *How an individual feels about the intervention* |
| Burden | *The perceived amount of effort that is required to participate in the intervention* |
| Ethicality | *The extent to which the intervention is a good fit with the individual's value system* |
| Intervention coherence | *The extent to which the participant understands the intervention and how it works* |
| Opportunity costs | *The extent to which benefits, profits or values must be given up to engage in the intervention* |
| Perceived effectiveness | *The extent to which the intervention is perceived as likely to achieve its purpose* |
| Self-efficacy | *The participant's confidence that they can perform the behaviour(s) required to participate in the intervention* |

#### Template two – identifying sub-themes

| **Main themes** | **Sub-themes** |
|---|---|
| Affective Attitude | A feeling of belonging<br>Relaxed and accepting atmosphere<br>Remote delivery suits pandemic, but you lose something<br>Safety and comfort |
| Burden | Structure<br>Experiencing emotions |

| | |
|---|---|
| | Homework can be burdensome<br>Remote access |
| Ethicality | Giving back<br>Trust |
| Intervention coherence | Handbook supports recall<br>Facilitation increases understanding<br>Accessibility aids<br>Tool-kit approach<br>Homework supports integration into life |
| Opportunity costs | Ease of access<br>Time |
| Perceived effectiveness | Handbook increases access to content<br>Stroke survivors learn from each other<br>Responsive facilitation<br>Need for additional support<br>Reviewing progress<br>Time post-stroke |
| Self-efficacy | Time to build relationships |

#### Template three – development of inductive main themes due to overlap in sub-themes across the TFA components

| **Sub-themes** | **Inductive main themes** |
|---|---|
| Homework supports integration into life<br>Time post-stroke<br>Tool-kit approach<br>Need for additional support | Engaging with ACT and applying it to life |
| Giving back<br>Trust<br>A feeling of belonging | Seeking community and support |

| | |
|---|---|
| Remote delivery suits pandemic, but you lose something<br>Stroke survivors learn from each other<br>Relaxed and accepting atmosphere<br>Time to build relationships | |
| Handbook supports recall<br>Handbook increases access to content<br>Accessibility aids<br>Safety and comfort<br>Ease of access<br>Remote access<br>Time post-stroke | Adaptations are important to support accessibility |
| Structure<br>Time<br>Homework can be burdensome<br>Reviewing progress | Intervention structure needs to be clear and account for burden |
| Safety and comfort<br>Trust<br>Responsive facilitation<br>Facilitation increases understanding<br>Need for additional support<br>Experiencing emotions | Facilitation supported learning and group cohesion |
| Need for additional support<br>Time to build relationships<br>A feeling of belonging | Moving on: what next after WAterS? |

#### Final template

| **Main themes** |
|---|
| Engaging with ACT and applying it to life |
| Seeking community and support |
| Adaptations are important to support accessibility |
| Intervention structure needs to be clear and account for burden |
| Facilitation supported learning and group cohesion |
| Moving on: what next after WAterS? |
